# Cardiovascular disease risk score derivation and validation in Abu Dhabi, United Arab Emirates. Retrospective Cohort Study

**DOI:** 10.1101/2024.03.19.24304561

**Authors:** Latifa Baynouna AlKetbi, Nico Nagelkerke, Ahmed Humaid, Noura AlAlawi, Rudina AlKetbi, Hamda Aleissaee, Noura AlShamsi, Hanan Abdulbaqi, Toqa Fahmawee, Basil AlHashaikeh, Muna AlDobaee, Mariam AlShamsi, Nayla AlAhbabi, AlYazia AlAzeezi, Fatima Shuaib, Jawaher Alnuaimi, Esraa Mahmoud, Alreem AlDhaheri, Mohammed AlMansoori, Sanaa AlKalbani, Wesayef AlDerie, Ekram Saeed, Nouf AlMarzooqi, Ahmed AlHassani, Amira AlAhmadi, Mohammed Sahyoni, Farah AlFahmawi, Ali AlAlawi, Yusra Sahalu, Aysha AYahyaee, Zinab AlAnsari, Khadija Doucoure, Rawan Ashoor, Reem AlShamsi, Maha AlAzeezi, Fatima AlMeqbaali, Noor Yahya, Shamma AlAlawi, Fatima AlKetbi

**Author notes:** Corresponding Author: Latifa Baynouna AlKetbi, MBBS, PhD, ArBFM, Ambulatory Healthcare Services, Al Ain, UAE.

## Abstract

Cardiovascular disease (CVD) risk assessment is key to rational decision-making in primary prevention. The CVD risk depends on dynamic factors requiring continuous equation updates.

**Design:** The Abu Dhabi Risk Study (ADRS) is the first and longest-duration retrospective cohort study in Abu Dhabi and the United Arab Emirates (UAE), aiming to develop 10-year risk prediction equations for CAD, stroke, and ASCVD (Atherosclerotic Cardiovascular Disease, combining CAD and stroke) and validate international risk equations.

**Method:** The included 8699 subjects are participants of the national cardiovascular screening program of 2011-2013 with an average follow-up of 9.2 years. They were assessed retrospectively in 2023 for health outcomes. The validation cohort, 2554 subjects, is another community-based screening program done in Abu Dhabi in the period from 2016 and 2017. With an average follow-up of 6.67 years.

**Results:** Of 8504 who were ASCVD-free, 250 had new coronary artery events. Identified risk factors for ASCVD in this population were the conventional risk factors such as age, gender, smoking, high cholesterol/HDL ratio, and diabetes diagnosis, in addition to low vitamin D level, and low glomerular filtration rate (GFR) levels.

Three ADRS prediction models were derived from Cox regression. The ADRS-CAD had a C-statistic of 0.899 compared to 0.828 of FRS (Framingham score) in the same population.

ADRS-Stroke had a c-statistic of 0.904. The ADRS-ASCVD had a c-statistic of 0.898 compared to 0.891 of PCE (pooled cohort equations) and 0.825 of FRS-CVD.

Applying the developed formulas to the validation cohort showed good predictability of CAD and ASCVD events with an ASCVD c-statistic of 0.825, for CAD the c-statistic was 0.799, and for stroke, it was 0.761. The PCE showed similar performance in this cohort with a c-statistic for ASCVD of 0.824.

**Conclusion:** This study demonstrated the value of tailoring risk assessments to local populations and healthcare contexts.

## Background

The age-adjusted mortality rate from heart disease (HD) and stroke decreased between 2011 and 2019 (1). However, the absolute number of deaths due to HD and stroke increased in association with the rapid growth of the population aged 65 years and older (1). In the UAE, cardiovascular diseases are the leading cause of death, accounting for 36.7% of all deaths, i.e., twice as many as the percentage of deaths attributed to cancer (2). Cardiovascular disease (CVD) risk assessment guidelines have utilized cardiovascular risk prediction equations to support clinical decision-making. These prediction equations are derived from multiple cohorts from different countries at different times. This “global approach” has raised concerns about their relevance in specific groups, including those defined by ethnicity or those with co- morbidities. In fact, a recent systematic review and meta-analysis found considerable heterogeneity in risk among studies and populations (3). A clear example is a U.S. cohort study of veterans with diabetes mellitus without prior ASCVD that concluded that existing ASCVD risk equations overestimate risk in these veterans, potentially impacting guidelines on statin therapy. Most importantly, it suggested that risk assessment can be improved by including several diabetes mellitus–related variables (4).

Despite these limitations, Cardiovascular disease (CVD) risk assessment guidelines are becoming an essential tool in individual patient care planning and in aiding policies on care for patients at high risk for cardiovascular diseases. The effectiveness of these CVD risk assessment equations in clinical practice was explored in a recent Cochrane systematic review on the use of short-term CVD risk estimation scores in primary prevention, which included 41 randomized controlled trials. There was significant heterogeneity and low-quality evidence for all of the individual trial results. It concluded that compared with usual care, providing quantitative CVD risk score information to clinicians and patients had a modest but statistically significant effect on levels of CVD risk factors and patients’ subsequent estimated 10-year CVD risk at follow-up. Providing risk information also led to increased initiation or intensification of lipid-lowering and antihypertensive medications. There was also evidence that harm resulting from quantitative risk assessments is unlikely (5). Therefore, risk assessment in the primary prevention of atherosclerotic cardiovascular diseases is highly recommended to start a process of shared decision-making between clinicians and patients with an emphasis on approaches to refine individual risk reductions for patients (6, 7).

Well-studied equations are the Framingham and the Pooled Cohort equations (8–10). The American College of Cardiology (ACC)/ AHA Pooled Cohort equations (PCEs) were derived from 5 community-based cohorts (ARIC [Atherosclerosis Risk in Communities]; CHS [Cardiovascular Health Study]; CARDIA [Coronary Artery Risk Development in Young Adults]; FHS [Framingham Heart Study]; FOS [Framingham Offspring Study]) (10). Two FRS equations were used, one for hard CAD, FRS, and one for ASCVD, FRS-CVD (8). The PCE has been widely validated and is broadly considered useful for the general US clinical population, which is reflected in the ACC/AHA clinical practice guidelines that recommend the use of the PCE for decision-making in primary prevention of ASCVD. The latter is also implemented in the Abu Dhabi ambulatory healthcare services for cardiovascular risk assessment. Local efforts to validate these equations were limited to one study of a small cohort extracted from hospital databases (11). Despite its limitations, this study gave early insights into additional risk factors for the incidence of cardiovascular diseases in the UAE, such as a lower globular filtration rate (GFR). In addition, a possible underestimation or overestimation of the risk in the UAE population may be caused by factors such as free access to healthcare with preventive healthcare services, or by ethnic differences. This study’s dual aims are the development of Abu Dhabi population-specific cardiovascular risk assessment equations and the validation of the Framingham and PCE equations in this population.

## Methods

This is a retrospective cohort study carried out in the Emirate of Abu Dhabi, United Arab Emirates. This cohort consists of UAE nationals who participated in the first Abu Dhabi population-wide cardiovascular screening program for adults aged 18 years or older, Weqaya, between the years 2010 and 2013. Participants were (re)assessed in 2023 with respect to changes in their health. Weqaya is a national screening program that started in Abu Dhabi in April 2008. All adults aged 18 years and above seeking to enroll in the UAE government’s free, comprehensive health insurance plan were required to participate in this screening program. It consisted of self-reported indicators, anthropometric measures, and hematological parameters. The Health Authority of Abu Dhabi oversees the Weqaya program, which was carried out by the Abu Dhabi Health Services Company through its Ambulatory healthcare services centers. Details of the Weqaya program are described elsewhere (12).

Data collected at baseline included demographic data. Self-reported health indicators included smoking status, physical activity, preexisting CVD (angina, heart attack, transient ischemic attack, stroke, other circulatory disorder), family history of premature cardiovascular disease (a first-degree relative with a heart attack or stroke before the age of 50 years), history of cardiovascular risk factors for diabetes, hypertension, and dyslipidemia and whether participants were taking medication for these conditions. Anthropometric measures included waist and hip circumference, body mass index (BMI in kg/m^2^), and a single arterial blood pressure reading. A digital automatic blood pressure monitor was used to measure blood pressure from the left arm with the patient relaxed and seated. Hematological parameters included non-fasting glucose (mmol/L), total cholesterol, high-density lipoprotein (HDL) cholesterol (mmol/L), glycosylated hemoglobin (HbA1c), vitamin D, and creatinine (12). We selected a time interval in Weqaya screening when vitamin D and creatinine were gathered for around twelve thousand participants. As there was missing data from some subjects, only subjects with completed data were included. The glomerular filtration rate was calculated based on the Chronic Kidney Disease Epidemiology Collaboration (CKD-EPI) equation (13).

Outcome assessment was done in 2023, the average follow-up period was 9.2, minimum follow-up of less than a year and a maximum of 12 years, Appendix 1. Data were collected by physicians and nurses who reviewed the Electronic Medical Records (EMR) of all subjects. As some death certification was documented in a different electronic system, the cause of death could only be determined if it was documented in the EMR. Therefore, some of the fatal cardiac events may be missing. In addition, there were some inaccuracies in the cause of death codes, mostly for deaths occurring in government hospitals or at home.

## Validation

The validation cohort is data from a community-based screening program done in AHS in the period from 2016 and 2017. The screening was comprehensive involving cardiovascular diseases and cancer. Subjects with complete data were included. Equal numbers of females and males were taken from the sample of the subjects who did the screening and who were free from CAD or ASCVD at baseline. The total sample of the cohort included was 2554, 1269 (49.7%) females and 1285 (50.3%) males. Outcomes were new acute coronary artery disease, diagnosis of coronary artery disease, or stroke. The outcome data were extracted in a report from the EMR. The average follow-up period for this cohort was 6.75 minimum less than a year and maximum of 8 years.

### Statistical analysis

Prediction models for CVD were developed using Cox survival regression with hard disease events as endpoints, both CAD and ASCVD (8, 14). These models were compared to the performance of the Framingham Risk Score (FRS) and PCE risk scores. ROC curves and c-statistics were used to evaluate the performance of the Cox-derived risk scores over a ten-year follow-up. Although all participants were included in the analysis, exclusion was done according to the criteria for PCE as per the published formula (9, 14).

### Developed formulas

Derived equations are shown in Appendix 2.

## Results

A total of 9579, 4886 females and 4693 males were eligible for inclusion. After performing stratified sampling based on gender, the study included a random sample of 8699: 4338 (49.9%) females and 4361 (50.1%) males, representing 5% of the Abu Dhabi population who underwent the baseline assessment.

In this cohort of 8699 subjects, 195 (2.2%) had previous coronary artery disease (CHD), 57 (1.3%) females and 138 (3.2%) males. When adding stroke patients (representing the ASCVD category), 343 (3.9%) had ASCVD before the screening date, 129 (3.0%) females and 214 (4.9%) males. There were also 243 pregnant women who were excluded from both analyses. Therefore, there were 8261, 4038 (48.9%) females and 4223 (51.1%) males included in the CAD cohort and 8123 included in the ASCVD cohort, 3976 (48.9%) females and 4147 (51.1%) males. Two equations were developed, one to predict hard coronary artery diseases (CAD) where only CAD-free patients were included and one to predict Athero-Sclerotic Cardiovascular diseases (ASCVD) where only ASCCVD-free patients were included.

The average follow-up years was 9.2 years (range 0- 12). Ten Percent of the subjects had a follow-up time of 2 years or less, while 75% of the patients were followed for nine years or more. Such long follow-up duration was facilitated by the integrated EMR system and the Abu Dhabi COVID-19 vaccination strategy. The population of Abu Dhabi was more than 97.97% vaccinated for COVID-19(15) and all vaccination visits required comprehensive chronic disease history and a physical examination, including BMI and blood pressure and sometimes repetition of some laboratory tests such as HA1C and kidney function. Only 18.4% of participants had their last (EMR) visit in 2019 or earlier. That increased to 21.5% in 2020, then 29.4% in 2021 and 53% in 2022, reaching 100% in 2023. In 2023 3968 (46.9%) still had EMR entries (supplement 1). The Cohort was followed up for mortality and other health conditions, myocardial infarction (MI), and stroke incidence until 2023.

From the 8261 CAD-free subjects, 250 (3%) developed coronary artery events during follow-up, 46 (1.1%) in females and 204 (4.7%) in males. The Framingham Risk Score FRS average was 2.69, 0.96 in females, and 4.44 in males, thus expecting 216 events, 38 in females, and 178 in males. Thus observed were 121,1% of the expected among females and 114.6% of the expected among males. The mean age of the first CAD was 63.3 years in males and 66.8 years in females. At the time of first CAD, 10% of the females were 49.6 years or younger, 10% of the males were 46.2 or younger, and 25% of females were 59.5 years or younger compared to 55 years or younger in males.

With regards to ASCVD, from the 8123 ASCVD-free subjects, 282 (3.5%) developed coronary artery events during the follow-up years. The incidence of CAD was 61 (21.6%) in females and 221 (78.4%) in males. The PCE average was 3.3, 1.3 in females, and 5.2 in males, expecting 272 events, 52 in females, and 220 in males, close to the reported number of events. The mean age of the first CAD was 63.3 years in males and 66.8 years in females. At the time of the events 10% of the females were 49.6 years or younger, 10% of the males were 46.2 or younger, and 25% of females were 59.5 years or younger compared to 55 years or younger in males.

Table 1 The age of the first CAD was 63.3 years in males and 66.8 years in females; 10% of the females were 49.6 years or younger, 10% of the males were 46.2 or younger, and 25% of females were 59.5 years or younger compared to 55 years or younger in males.

**Table 1.** Characteristics of the development and validation cohorts’ subjects distributed by sex.

|  |  | Development Cohort |  |  |  | Validation Cohort |  |
| --- | --- | --- | --- | --- | --- | --- | --- |
|  |  | Subjects with no ASCVD at baseline |  | Subjects with no CAD at baseline |  | Subjects with no ASCVD or CAD at baseline |  |
|  |  | Mean | SD | Mean | SD | Mean | SD |
| Female | Age (years) | 37.6 | 14.1 | 37.7 | 14.1 | 46.5 | 12.8 |
|  | MEAN BP | 85.8 | 10.5 | 85.8 | 10.5 | 90.2 | 11.8 |
|  | SBP | 116.3 | 15 | 116.3 | 15 | 124.4 | 17.5 |
|  | DBP | 70.6 | 10 | 70.6 | 9.9 | 73.1 | 11.2 |
|  | Random Glucose | 5.8 | 2.2 | 5.8 | 2.2 | 4.9 | 3.4 |
|  | Total Cholesterol | 4.8 | 1 | 4.8 | 1 | 4.8 | 0.9 |
|  | HBA1C | 5.7 | 1 | 5.7 | 1 | 5.9 | 1.1 |
|  | HDL | 1.4 | 0.3 | 1.4 | 0.3 | 1.3 | 0.3 |
|  | Vitamin D | 34.6 | 22.7 | 34.7 | 22.7 | 64.4 | 30.1 |
|  | BMI | 29 | 6.8 | 29 | 6.7 | 30.7 | 6.2 |
|  | Waist Hip Ratio | 0.9 | 0.1 | 0.9 | 0.1 |  |  |
|  | Cholesterol HDL ratio | 3.6 | 1.1 | 3.6 | 1.1 | 3.6 | 1.07 |
|  | Deviation from mean GFR | 2.6 | 17.2 | 2.6 | 17.2 | 0.39 | 14.9 |
|  | GFR | 114 | 17.2 | 113.9 | 17.3 | 106.9 | 14.9 |
| Male | Age (years) | 39.6 | 15.4 | 39.6 | 15.5 | 42.4 | 12.3 |
|  | MEAN BP | 92.7 | 10.2 | 92.7 | 10.3 | 92.3 | 10.1 |
|  | SBP | 124.7 | 13.8 | 124.8 | 13.9 | 125.7 | 13.1 |
|  | DBP | 76.6 | 10.1 | 76.7 | 10.1 | 76.5 | 10.3 |
|  | Random Glucose | 6.4 | 3 | 6.4 | 3 | 4.4 | 3.6 |
|  | Total Cholesterol | 4.8 | 1.1 | 4.8 | 1.1 | 4.8 | 0.98 |
|  | HBA1C | 5.9 | 1.2 | 5.9 | 1.2 | 5.8 | 0.97 |
|  | HDL | 1.1 | 0.3 | 1.1 | 0.3 | 1.1 | 0.2 |
|  | Vitamin D | 36.4 | 20.9 | 36.5 | 21 | 59.9 | 28.9 |
|  | BMI | 28.5 | 5.8 | 28.5 | 5.8 | 29 | 5.77 |
|  | Waist Hip Ratio | 0.9 | 0.1 | 0.9 | 0.1 |  |  |
|  | Cholesterol HDL ratio | 4.5 | 1.8 | 4.5 | 1.8 | 4.5 | 1.3 |
|  | Deviation from mean GFR | -2.5 | 18 | -2.5 | 18 | -0.39 | 18.8 |
|  | GFR | 108.9 | 18 | 108.7 | 18.3 | 106.1 | 18.8 |
|  |  | **Subjects with no ASCVD at baseline** |  | **Subjects with no CAD at baseline** |  | **Subjects with no ASCVD or CAD at baseline** |  |
|  |  | Number | % | Number | % | Number | % |
| Female | On Hypertension treatment | 452 | 11.40% | 466 | 11.50% | 339 | 26.70% |
|  | Family History of Heart attack | 193 | 4.90% | 196 | 4.90% |  |  |
|  | Family History of stroke | 71 | 1.80% | 73 | 1.80% |  |  |
|  | History of walking adequately | 2243 | 58.40% | 2277 | 58.30% |  |  |
|  | Walking status | 2243 | 58.40% | 2277 | 58.30% |  |  |
|  | Chronic Kidney Disease | 36 | 0.90% | 38 | 0.90% | 12 | 0.90% |
|  | Cancer | 39 | 1.00% | 40 | 1.00% |  |  |
|  | Hypertension | 597 | 15.00% | 616 | 15.30% | 379 | 29.90% |
|  | Diabetes Mellitus | 727 | 16.90% | 745 | 18.40% | 336 | 26.50% |
|  | Current Smoker | 18 | 0.50% | 19 | 0.50% | 1 | 0.10% |
|  | Dyslipidemia before | 671 | 16.90% | 690 | 17.10% | 235 | 18.50% |
|  | On statin | 535 | 13.6% | 550 | 13.8% | 536 | 42.2% |
| MALE | On Hypertension treatment | 598 | 14.40% | 619 | 14.70% | 292 | 22.70% |
|  | Family History of Heart attack | 143 | 3.50% | 148 | 3.50% |  |  |
|  | Family History of stroke | 66 | 1.60% | 67 | 1.60% |  |  |
|  | Walking status | 2836 | 70.90% | 2881 | 70.70% |  |  |
|  | Chronic Kidney Disease | 46 | 1.10% | 52 | 1.20% | 11 | 0.90% |
|  | Cancer | 23 | 0.60% | 24 | 0.60% |  |  |
|  | Hypertension | 868 | 20.90% | 899 | 21.30% | 332 | 25.80% |
|  | Diabetes Mellitus | 996 | 19.10% | 1027 | 24.30% | 231 | 18.00% |
|  | Current Smoker | 687 | 16.60% | 695 | 16.50% | 158 | 12.30% |
|  | Dyslipidemia before | 794 | 19.10% | 823 | 19.50% | 230 | 17.90% |
|  | On statin | 647 | 16.1% | 673 | 16.4% | 471 | 36.7% |

This cohort is relatively young, in both the ASCVD and CAD cohorts, with mean ages of 37.6 in females and 39.6 in males in the ASCVD cohort and 37.7 in females and 39.6 in males in the CAD cohort. Males had a higher blood pressure average than females and a higher prevalence of hypertension, dyslipidemia, and diabetes mellitus than females despite being more physically active than females. Around 16% of both CAD and ASCVD cohorts reported smoking, almost all males. Males have lower GFR than females, with an average of -2.5 less than the mean of the cohort compared to +2.6 in females in both cohorts. Table 1 Using Cox regression, significant predictors of new-onset CHD events in the follow-up years from risk factors assessed at baseline in 2011-2013 included all traditional risk factors in the Framingham equation as listed in Table 2, shown with regression coefficients and hazard ratio. In addition, there were two new risk factors unique to this cohort: lower vitamin D levels and lower GFR levels. Regarding the traditional risk factors, older age increased the risk of first CAD by 6.9% for each additional year, while male sex increased risk 8.5 fold. Diabetes was a major predictor that increased risk 4.7 fold. If diabetes co-existed with lower GFR the risk increased by 2% for each. History of hypertension and higher blood pressure levels increased risk. The risk increased by 39% for hypertension diagnosis, and Table 2 shows the HR for different blood pressure categories of subjects. There was a 6.9% increased risk for each unit increase in Cholesterol/HDL ratio. Smoking among males increased the risk by 73%. Lower vitamin D level was associated with increased risk by 7% for each ng/ml decrease in Vitamin D. Low GFR was another new independent risk factor. GFR also had a significant interaction with sex and with a diagnosis of diabetes. In females, the effect of these risk factors appeared to be stronger than in males.

**Table 2.** Cox regression for determinants of the new incidence of A. Hard Coronary Artery Diseases in the cohort, B. Strock, C. ASCVD, D.Death among the cohort subjects. Only significant variables are listed.

| **A. Cox regression for determinants of new incidence of Hard Coronary Artery Diseases (CAD) in the cohort** |  |  |  |  |  |
| --- | --- | --- | --- | --- | --- |
|  | **B** | **P value** | **HR** | **HR 95.0% CI** |  |
| Vitamin D (ng/ml) | -0.007 | 0.033 | 0.993 | 0.986 | 0.999 |
| Deviation from cohort GFR mean (ml/min/1.73 m2) | -0.039 | <.001 | 0.962 | 0.946 | 0.978 |
| Sex (0 female and 1 male) | 2.136 | <.001 | 8.463 | 4.042 | 17.72 |
| Current Smoker (yes =1) | 0.55 | 0.002 | 1.733 | 1.23 | 2.441 |
| Age (year) | 0.067 | <.001 | 1.069 | 1.055 | 1.082 |
| Hypertension | 0.327 | 0.039 | 1.387 | 1.017 | 1.89 |
| Cholesterol/HDL ratio (mmol/L) | 0.067 | 0.046 | 1.069 | 1.001 | 1.141 |
| Diabetes Mellitus | 1.543 | <.001 | 4.678 | 2.138 | 10.235 |
| BP stages |  | 0.042 |  |  |  |
| BP Normal: Below 120/80 (1) | -1.118 | 0.036 | 0.327 | 0.115 | 0.931 |
| BP Elevated: 120 to 129/Less Than 80 (2) | -1.148 | 0.03 | 0.317 | 0.112 | 0.897 |
| BP Stage 1 High Blood Pressure: 130 to 139/80 to 89 (3) | -1.378 | 0.009 | 0.252 | 0.089 | 0.712 |
| BP Stage 2 High Blood Pressure: 140 and Above/90 and Above (4) | -1.093 | 0.038 | 0.335 | 0.12 | 0.939 |
| BP Hypertension Crisis (5) | -0.698 | 0.21 | 0.498 | 0.167 | 1.483 |
| Deviation from cohort GFR mean (ml/min/1.73 m2)* Sex (0 female and 1 male) | 0.027 | <.001 | 1.028 | 1.012 | 1.044 |
| Diabetes Mellitus * Sex (0 female and 1 male) | -0.669 | 0.075 | 0.512 | 0.246 | 1.069 |
| Diabetes Mellitus * Deviation from cohort GFR mean (ml/min/1.73 m2) | 0.019 | 0.008 | 1.02 | 1.005 | 1.034 |
| **B. Cox regression for determinants of new stroke among the cohort subjects** |  |  |  |  |  |
|  | **B** | **P value** | **HR** | **HR 95.0% CI** |  |
| Age (year) | 0.084 | <0.001 | 1.088 | 1.067 | 1.11 |
| CurrentSmoker | 1.053 | 0.003 | 2.866 | 1.423 | 5.773 |
| SBP | 0.024 | 0.004 | 1.024 | 1.008 | 1.041 |
| Hypertension * Sex (0 female and 1 male) | -0.646 | 0.075 | 0.524 | 0.257 | 1.068 |
| Age (year) * Sex (0 female and 1 male) | 0.011 | 0.053 | 1.011 | 1 | 1.022 |
| **C. Cox regression for determinants of new incidence of Atherosclerotic Cardio Vascular Diseases (ASCVD) in the cohort** |  |  |  |  |  |
|  | **B** | **P value** | **HR** | **HR 95.0% CI** |  |
| Vitamin D (ng/ml) | -0.007 | 0.029 | 0.993 | 0.987 | 0.999 |
| Sex (0 female and 1 male) | 1.516 | <0.001 | 4.554 | 2.863 | 7.244 |
| Current Smoker | 0.611 | <0.001 | 1.841 | 1.348 | 2.515 |
| Diabetes Mellitus | 0.332 | 0.016 | 1.394 | 1.064 | 1.827 |
| On hypertension treatment | 0.305 | 0.025 | 1.357 | 1.039 | 1.773 |
| MEAN BP SBP/DBP<sup>2</sup> | 0.012 | 0.037 | 1.012 | 1.001 | 1.024 |
| Cholesterol/HDL ratio (mmol/L) | 0.068 | 0.049 | 1.07 | 1.07 | 1.145 |
| Age<sup>2</sup> (year) | -0.001 | <0.001 | 0.999 | 0.998 | 0.999 |
| Deviation from cohort GFR mean (ml/min/1.73 m<sup>2</sup>) | -0.029 | <0.001 | 0.971 | 0.959 | 0.983 |
| Age (year) | 0.204 | <0.001 | 1.226 | 1.148 | 1.309 |
| Deviation from cohort GFR mean (ml/min/1.73 m<sup>2</sup>) * Sex (0 female and 1 male) | 0.031 | <0.001 | 1.032 | 1.017 | 1.047 |
| **D. Cox regresion for determinants of new Death among the cohort subjects** |  |  |  |  |  |
|  | **B** | **P value** | **HR** | **HR 95.0% CI** |  |
| Age (year) | 0.057 | <.001 | 1.058 | 1.04 | 1.077 |
| Sex (0 female and 1 male) | 0.62 | 0.01 | 1.859 | 1.161 | 2.976 |
| Cancer history | 1.218 | 0.04 | 3.381 | 1.06 | 10.782 |
| Walking status (1 adequate 0 not adequate) | -0.386 | 0.098 | 0.68 | 0.431 | 1.074 |
| Use of Lipid Lowering Medications | -0.495 | 0.075 | 0.61 | 0.353 | 1.052 |
| Hypertension * Deviation from cohort GFR mean (ml/min/1.73 m<sup>2</sup>) | -0.021 | 0.004 | 0.979 | 0.965 | 0.993 |

C statistics for risk scores derived from the Cox regression analyses were used to assess the model and compare them to the FRS as in Figure 1. a. The c-statistics for the ADRS-CAD was 0.899 and for the FRS, 0.828. The maximum Kolmogorov-Smirnov (K-S) statistics for this model is 0.662 for the ADRS-CAD and 0.525 for the FRS. The sensitivity of the developed model at different cutoff points of 1, 4, 5, 7.5, 10, and 20 are listed in Figure 1; reasonable sensitivity and specificity are 4 and 5; 89.3 and 75.6, and 86.3 and 78.5, respectively. Moving towards higher cutoff points, improvement in specificity is associated with less sensitivity, with sensitivity reaching 53.2 at point 20.

**Figure 1.**
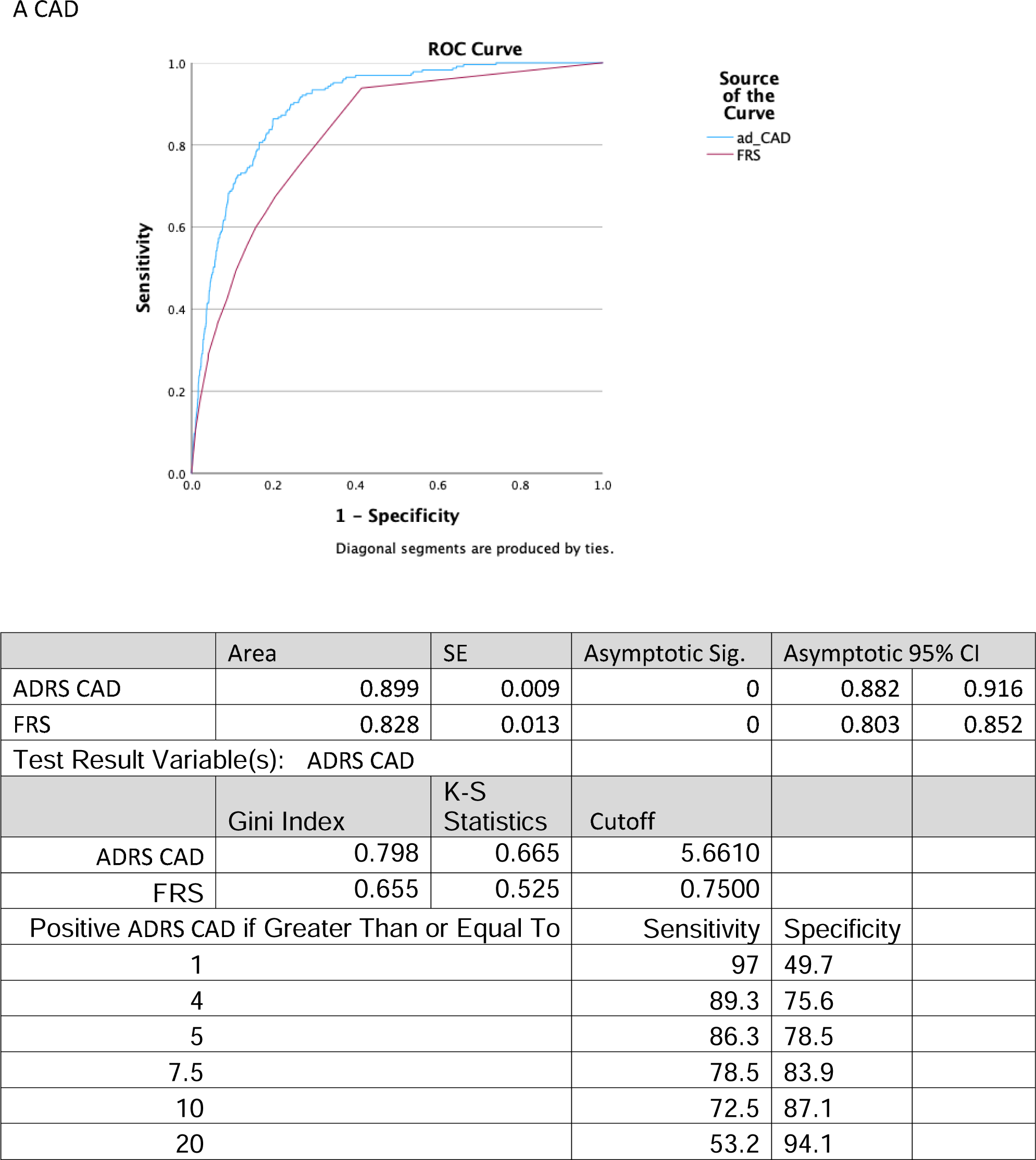
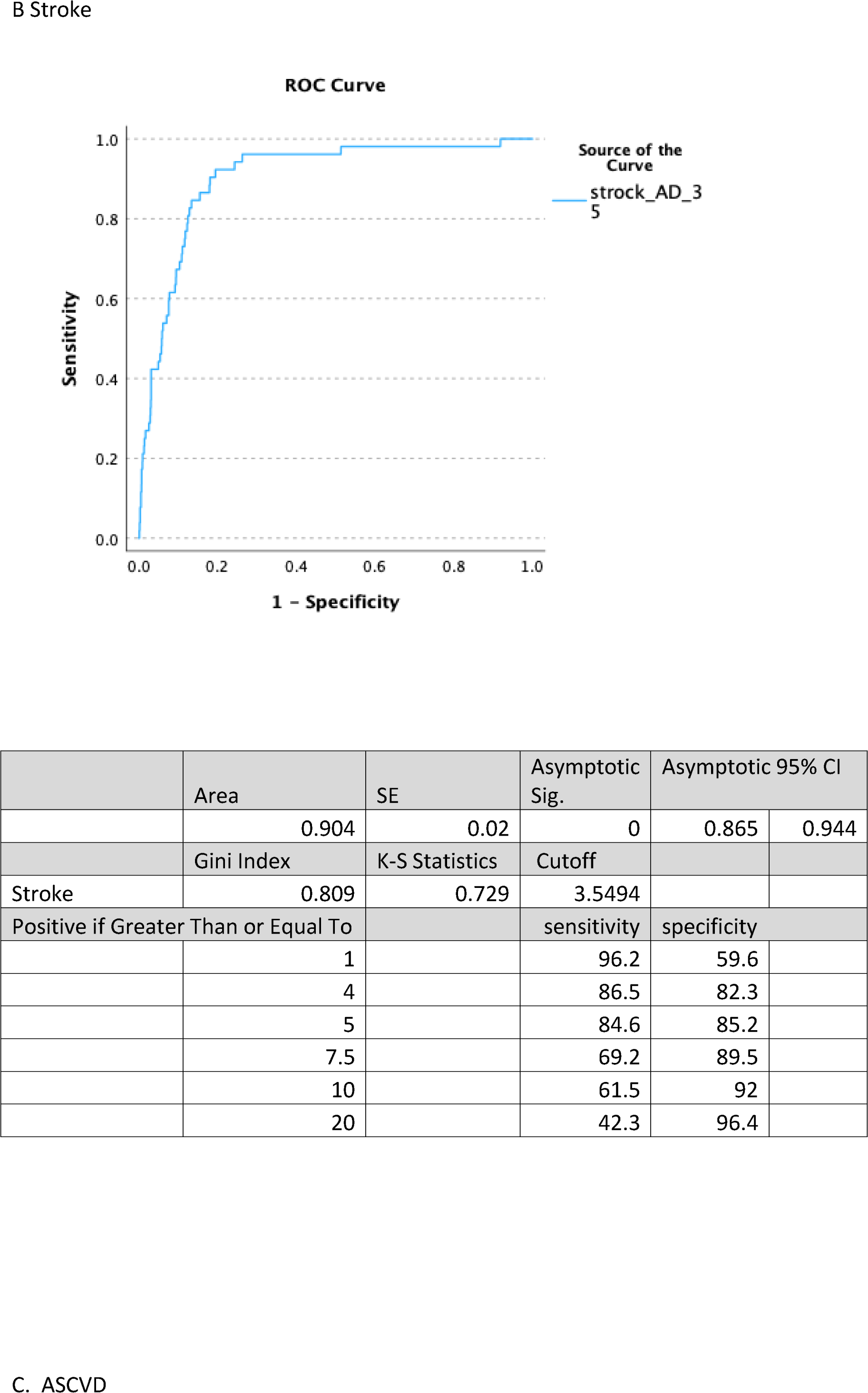
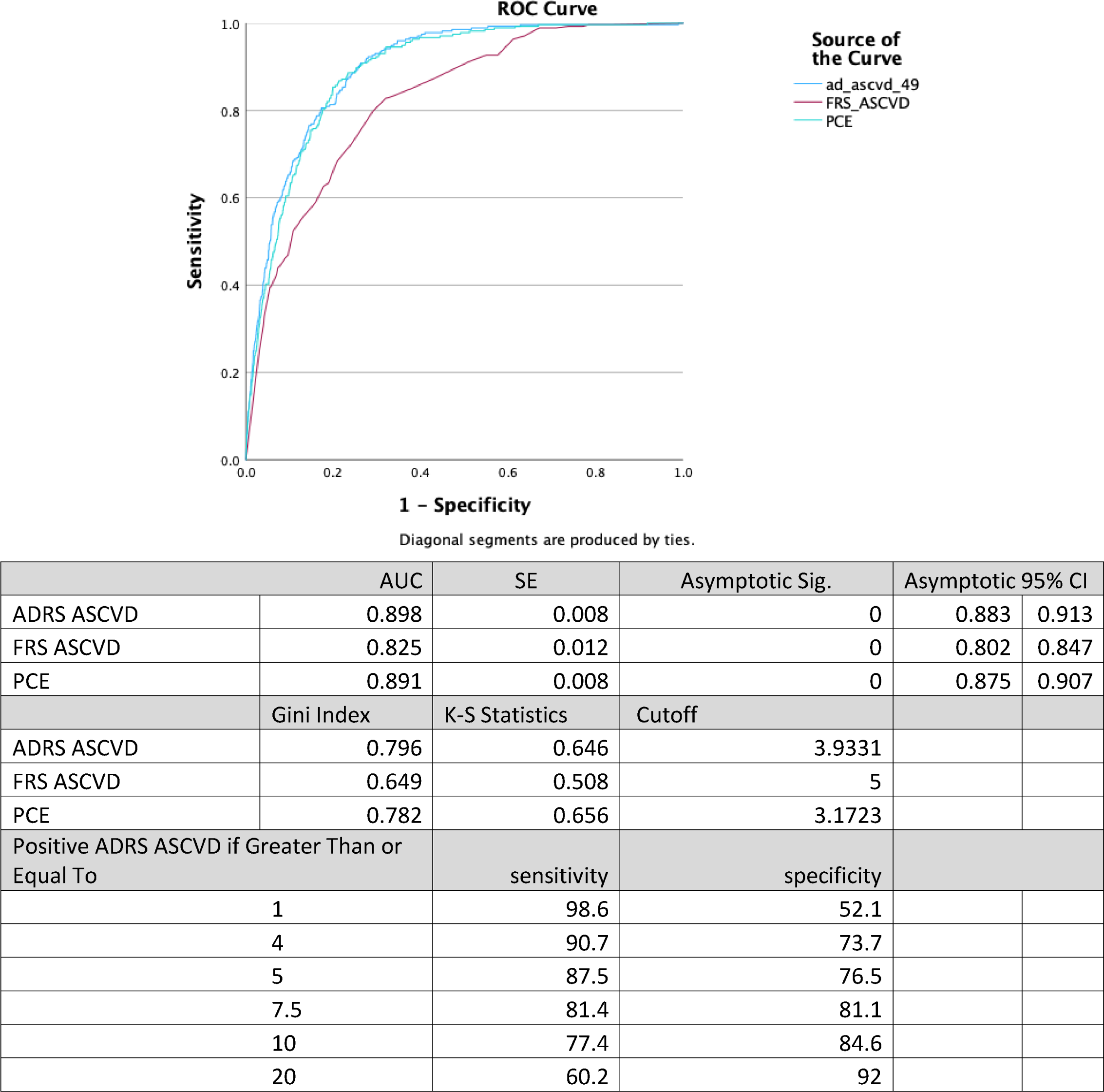
Area Under the Curve (AUC), c-statistic, with coordinates of the ROC curve noting the sensitivity and specificity of the equation at different cutoff points. A. The derivation cohort of CAD-free subjects followed for ASCVD events over the follow-up period B. The derivation cohort f stroke-free subjects followed for new stroke development over the follow-up period C. The derivation cohort of ASCVD-free subjects followed for ASCVD events over the follow-up period.

Determinants of stroke in this population are very different than those of CAD. Only older age, smoking history, raised SBP, and the interaction of hypertension and sex, hypertensive women being at a higher risk of stroke and the interaction of sex and age, with older men being at elevated risk. Table 2 B. Our score’s c statistic was excellent, 0.904 (0.865-0.944) with excellent fit, K-S Statistics 0.729. At a cutoff 5% or higher, hazard risk sensitivity is 84.6%, and Specificity is 85.2%.

With regards to ASCVD, a second equation was developed from Cox regression with ASCVD events as the outcome and baseline risk factors at screening time in 2011-2013 as the independent variables. Again, the same risk factors as in the PCE and FRS-CVD were identified. The hazard ratio (HR) for these risk factors is shown in Table 2C. Gender had the highest HR of 4.6, smoking nearly doubled the risk of ASCVD with an HR of 1.8 (1.3-2.5), and diabetes similarly increases the risk by 39%, HR=1.39. Older age was associated with a 15% increase in risk for each additional year. Higher blood pressure is associated with a higher risk of 1.2% for each mm Hg increase in mean blood pressure and 35% if the patient is treated for hypertension. High cholesterol/ HDL ratio is associated with increased risk as well, 7% for each unit increase. Regarding the two variables not included in FRS-CVD and PCE scores, for each nmol/L reduction in vitamin D, there is an increased risk of ASCVD of 0.7% p-value =0.029, HR=0.993(0.98-0.99). Similarly, one mL/min/1.73m2 GFR lower deviation from the mean increases the risk of ASCVD by 2.9 %, HR=0.971. There was a significant interaction between the deviation from GFR mean and sex, with males with lower GFR having higher risks of ASCVD.

C statistics derived from the Hazard ratio from the Cox regression analysis were used to assess the model and compare it to the PCE, FRS-CVD as in Figure 1.c. The c-statistics for the ADRS-ASCVD were 0.898, 0.891 for the PCE, and 0.825 for the FRS-CVD. The maximum Kolmogorov-Smirnov (K-S) statistics for this model are 0.646 for the ADRS-CAD, 0.656 for the PCE, and 0.508 for the FRS-CVD. The sensitivity of the developed model at different cutoff points of 1, 4, 5, 7.5, 10, and 20 are in Figure 1; reasonable sensitivity and specificity are 5 and 7.5, 87.5 and 76.5, and 81.4 and 81.1, respectively. Moving towards higher cutoff points, improvement in specificity is associated with less sensitivity, with sensitivity reaching 60.2 at 20.

The PCE, FRS-CVD equation performance is satisfactory in this population, but there appears to be a great mismatch between the ADRS-CAD and FRS-CVD. Both PCE and the ADRS performed very well in identifying high-risk patients and were superior to the FRS. The ADRS-CAD identified 183(73.2%) patients who experienced new CADs during follow-up as having 7.5% risks or higher, compared to 122 (48.8%) by the FRS. The ADRS-ASCVD identified 227 subjects (80.4%) who experienced an ASCVD during follow-up as high risk, i.e. risk score more or equal to 7.5, compared to 182 (64.5%) by the PCE and 192 (68.1%) by the FRS-CVD. Appendix 3A shows the performance of the ADRS and PCE in risk assessment.

Significant baseline risk factors for death as an outcome using Cox regression are shown in Table 2 d, with age, the interaction of hypertension diagnosis with lower deviation in GFR from the mean of this population cohort, cancer diagnosis, less physical activity, male sex and less treatment with LLMs. Surprisingly, the following were not significant predictors of death: CAD, stroke, smoking, and diabetes. Lipid-lowering medication (LLM) use was very common in this cohort, (table 3). At baseline, 15.1% of the cohort were on LLMs, 16.4% of males, and 13.8% of females. During the follow-up, the prevalence of use increased. Of patients who developed CAD later during the follow-up years, almost half, 49%, were using LLM at baseline, which increased to 92% during the follow-up years. In those who did not develop CAD, only 14% used LLM at baseline, and during follow-up, this increased to 29%. Interestingly, the use of LLM was positively correlated to the measured Hazard ratio categories as shown in Table 3, ranging from 2% in the lowest risk category to 92% in the highest. We explored the effect of LLMs in reducing cardiovascular risk and its outcome, mainly mortality. Figure 2 shows the Kaplan-Meier for survival (death rate) among patients for users and non-users of LLMs stratified by ADRS category. As shown, subjects on LLMs had better survival. Kaplan-Meier survival analysis shows the difference in survival of the subgroup of the cohort based on future development of either CAD or ASCVD stratified by ADRS risk categories. The mortality rate was higher beginning from year two among those who developed CAD and were not on LLMs. At year five, for those who developed CAD and were on LLMs, the mortality started to increase but less than for those who were not on LLMs, and the rise in mortality was mainly among the higher ADRS risk categories. The other two groups with no CAD developed no difference in survival between those on LLMs and those not on LLMs. Nevertheless, as in Figure 2, those who are on LLMs have higher ADRS-CAD / ADRS-ASCVD HR. Similarly, those who died were at higher risk of CAD or ASCVD, as ADRS HR was higher in Figure 2.

**Figure 2a.**
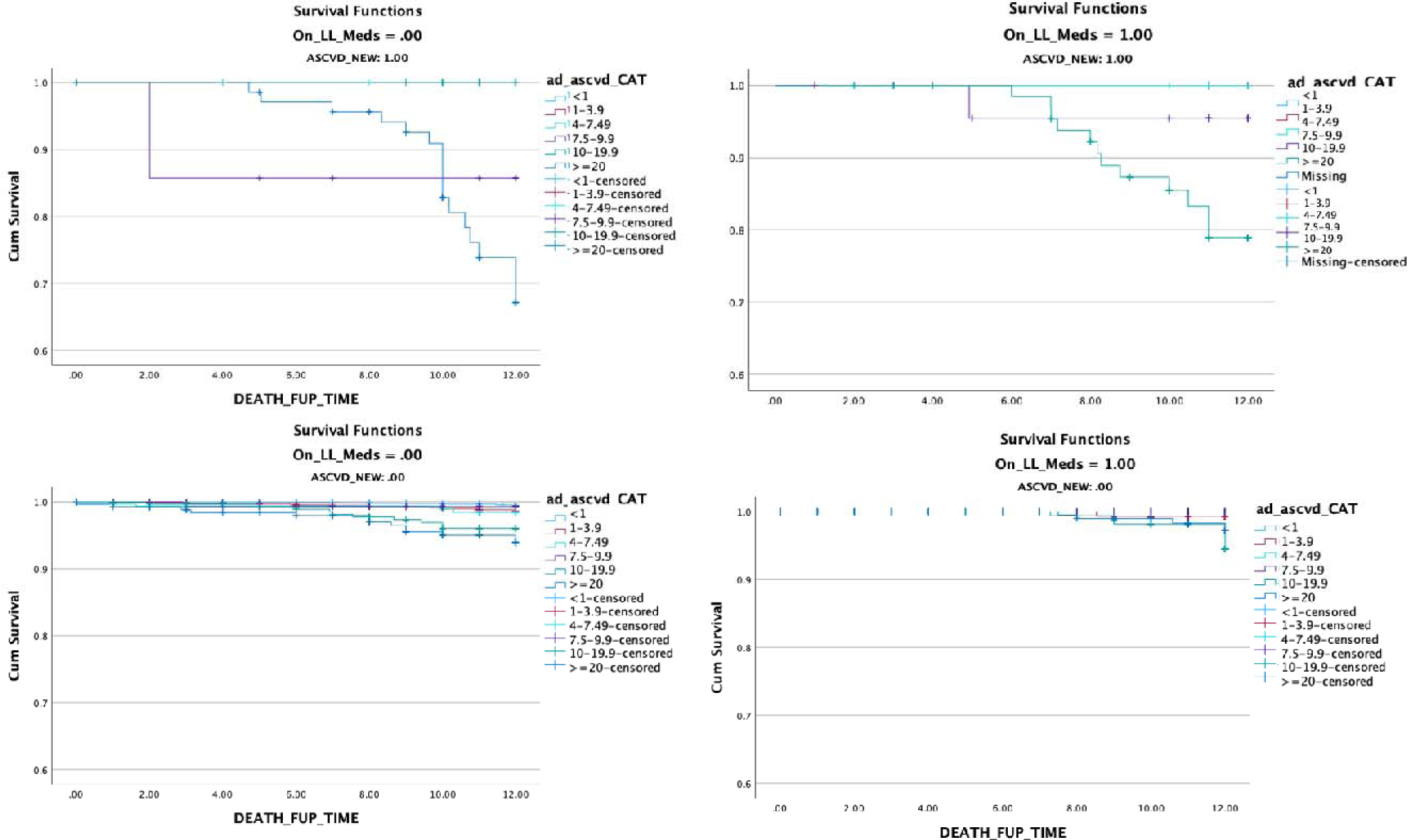
a Kaplan-Miere graph of mortality in two strata, on LLM and not on LLM, stratified into ADRS-ASCVD 5 risk levels; <1%, 1-3.99%, 4%-7.49%, 7.5-9.99%, 10-19.9%, and >=20

**Figure 2b.**
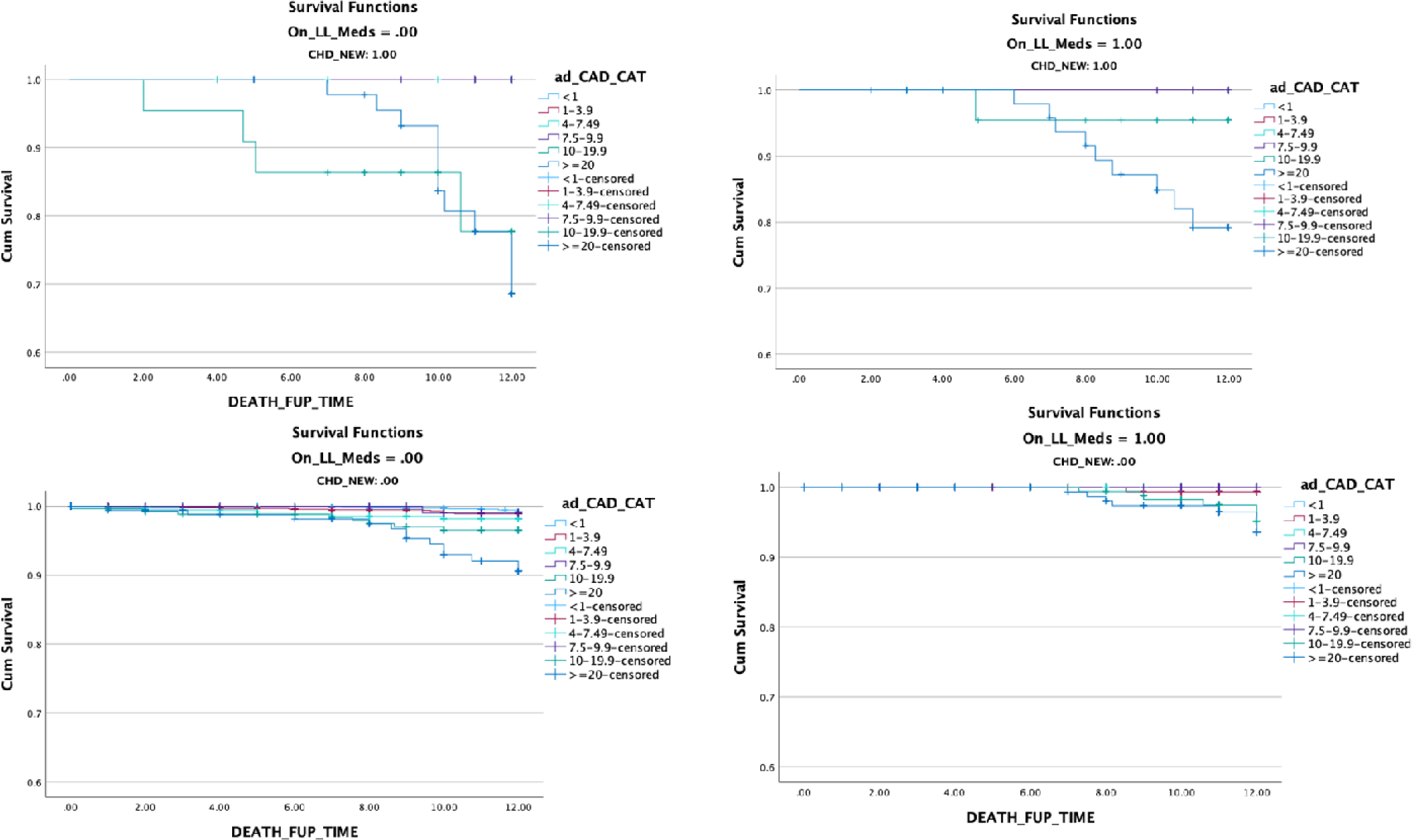
b Kaplan-Miere graph of mortality in two strata, on LLM and not on LLM stratified into ADRS-CAD 5 risk levels; <1%, 1-3.99%, 4%-7.49%, 7.5-9.99%, 10-19.9%, and >=20

With regards to mortality, in subjects with CAD or ASCVD during the follow-up years, one event (0.4%) was fatal; 4 (1.6%) were ASCVD-related, 18 (7.2%) occurred later with no documentation in the EMR of the death cause; 8 (3.2%) were unrelated to ASCVD. This attenuation of risk of death reflects what is concluded from Cox regression in Table 2c, that being on LLMs decreases the risk of death. As well in univariant logistic regression between death and duration of statin use, there is at least a 1.4% reduced risk of death for every year of statin use, noting the unknown compliance rate, B= - 0.157, P value = 0.001 and OR= 0.86 (0.78-0.94).

## Validation

Table 1 shows the characteristics of the subjects of the validation database. The average age for males was 42.4 years and for females 46.5 years. Prevalence of hypertension was higher in the validation cohort than in the derivation cohort, with 29.9% among females and 25.8% among males, similarly, diabetes was more prevalent in the validation cohort, 26.50% among females and 18% among males. More subjects were on statins at screening in the validation cohort, 39.4%. Vitamin D deficiency was higher in the validation cohort 64.4 in females and 59.9 in males compared to 34.7 and 36.4 respectively.

In the validation cohort, 352 ASCVD events occurred during the 6.7 years of follow-up period, 169 (13.3%) among females and 183 (14.2%) among males. Of those there were 301 CAD events, 144 (11.3%) among females and 157(12.2%) among males and 94 stroke new cases, 41 (3.2%) among females and 53 (4.1%) among males.

The developed formulae for the three outcomes were externally validated on the validation cohort to assess the prediction ability of the developed formula. For ASCVD the c-statistic was 0.825, for CAD the c-statistics was 0.799, and for stroke, it was 0.761. Figure 3 shows the Area Under the Curve (AUC), c-statistic, with coordinates of the ROC curve noting the sensitivity and specificity of the equation at different cutoff points. The three formulas performed well among the validation cohort as all c-statistics exceeded 0.7. The PCE showed similar performance in this cohort with a c-statistic for ASCVD of 0.824.

**Figure 3.**
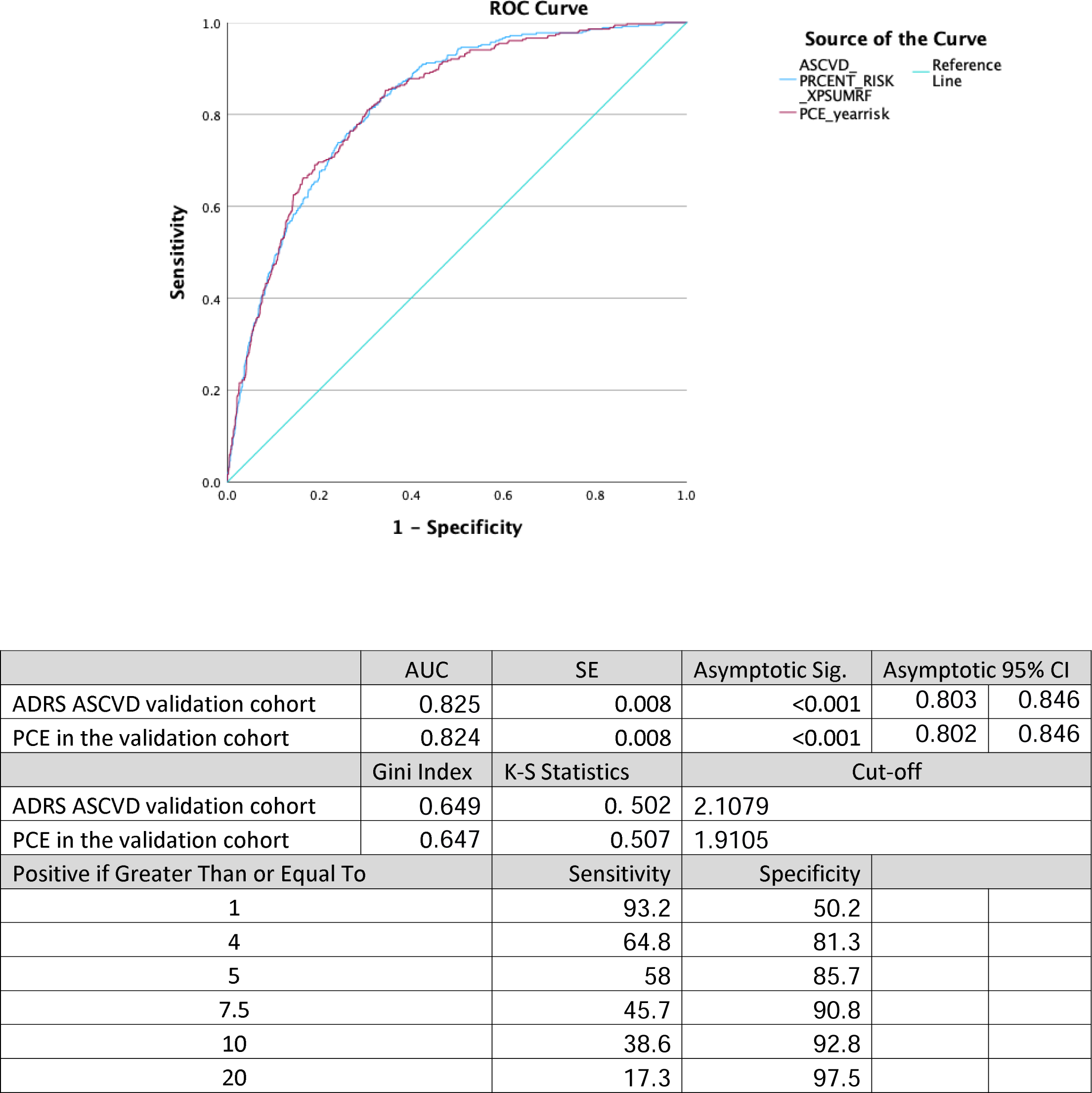
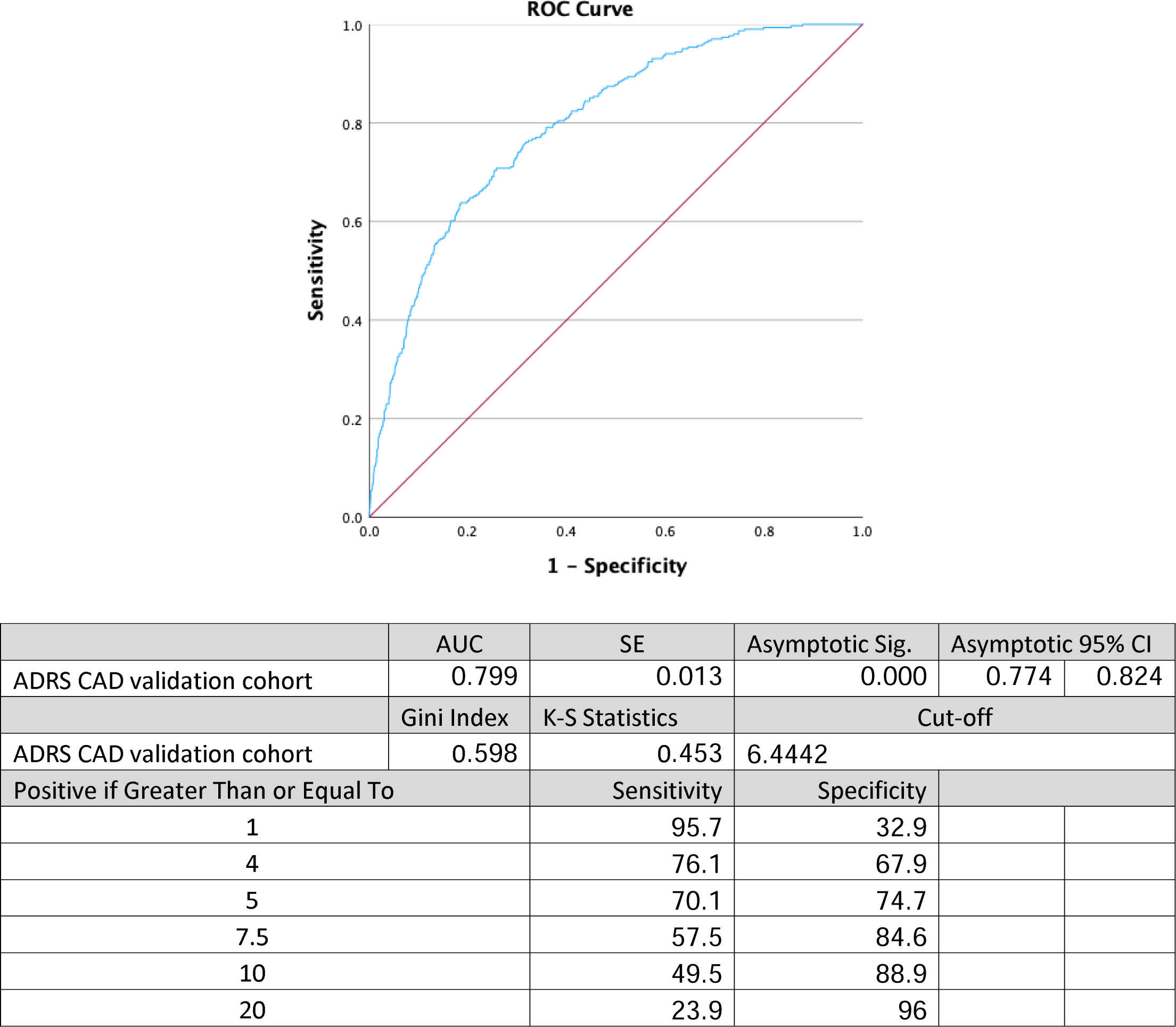
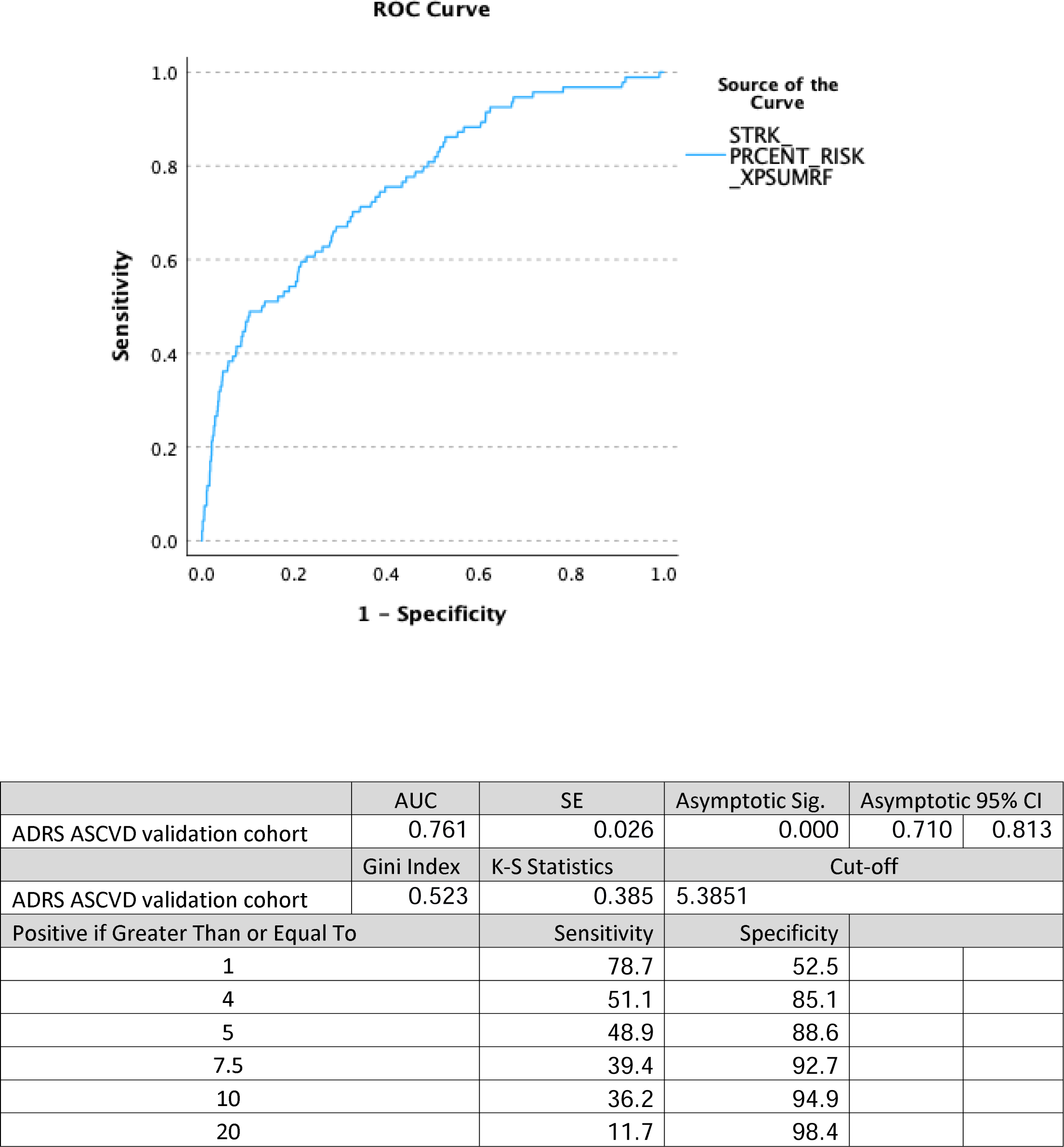
Area Under the Curve (AUC), c-statistic, with coordinates of the ROC curve noting the sensitivity and specificity of the equation at different cutoff points. A. The validation cohort of CAD-free subjects followed for ASCVD events over the follow-up period B. The validation cohort of ASCVD-free subjects followed for ASCVD events over the follow-up period. C. The validation cohort of Stroke-free subjects followed for stroke events over the follow-up period.

In this validation cohort’s subjects who developed an ASCVD, there is a similar number of patients identified to be low risk by the ADRS to the PCE in both the derivation and the validation cohorts. For example, there are 148 (42%) subjects with ASCVD identified by the ADRS-ASCVD as having a risk below 5% in the validation cohort and 144 (40.9%) patients by the PCE. In subjects who developed CAD, only 90 (29.9%) had a risk assessment of less than 5%, and in those who developed new stroke 48 (51%). Also, of subjects who developed ASCVD, 148 (42%) were identified to have a high risk, more or equal to 7.5%, by ADRS- ASCVD, and 192 (54.5%) by PCE were identified to have a higher risk at baseline.

In subjects with no ASCVD events developed during follow-up, 161 (45.7%) of them were identified as risk higher than 7.5% by the ADRS-ASCVD compared to 148 (42%) identified by the PCE. The identification of higher risk of CAD, those with a risk of 7.5% or more, was better with 173 (57.5%) patients of the 301 who developed CAD identified as high risk. In stroke, only 37 (39.4%) who had a stroke were in that category. Appendix 3 shows the stratification of the risk categories based on the ADRS formula in relation to the PCE risk categories and distributed by the occurrence of an ASCVD event during the follow-up period. In the derivation cohort in A, B, C, and the validation cohort in D. Overall, there is less agreement between risk categories of the ADRS and PCE in the moderate risk, 5% to 7.5% while in the lower and higher risk categories, the agreement is better.

## Discussion

This study developed equations for the prediction of the risk of CAD and ASCVD for the Abu Dhabi population that appear to have slightly better performance in this population than equations developed in other countries (16). Importantly, there are differences in risk factors as several new risk factors were identified, lower vitamin D and lower GFR levels, as well as their interaction with established risk factors, which, in addition to refining risk prediction, may also suggest novel avenues to risk reduction (17).

Such variability in risk factors and the availability of reliable and readily obtainable biomarkers for monitoring cardiovascular disease risk and, probably most importantly, the availability of new therapies with cardiovascular benefits and better side effects profiles justifies the continuous reevaluation of risk prediction scores (18).

In addition, we further refined the role of diabetes and how it interacts with other factors in the risk of CAD. Specifically, we found a positive interaction between diabetes and female sex. This interaction of sex and diabetes was also suggested by a systematic review of 64 cohorts; the hazard rate for CAD associated with diabetes was 2.82 in women and 2.16 in men (19). Such sex interaction with diabetes and the high prevalence of diabetes may contribute to the overall incidence of CAD in our population. The prevalence of over 20% (20) is very high compared to other countries where the FRS and PCE were developed, with a US prevalence of 8.9% (21). The prediction of risk may be influenced by such epidemiological differences.

With regards to vitamin D, 1,25(OH), 2D plays a pivotal role in adequate cardiac and vascular function (22). Markers of vitamin D metabolism vary significantly by race/ethnicity and are attributable, at least partly, to genetic ancestry (23). These differences likely may explain racial/ethnic differences in the risk and prevalence of vitamin D-related diseases (23). Its prevalence is <20% of the population in Northern Europe but up to 80% in Middle East countries (24). Nevertheless, there remains controversy in the literature on its impact on health outcomes (25). In a prospective population-based study in Lausanne, Switzerland, Patriota et al. reported a significant inverse association between vitamin D levels and CVD events but not with CVD or overall mortality (26). This is similar to the results of our study, as the significant association was only with cardiac events but not overall SCVD or mortality. By contrast, in a Mendelian randomization analysis from the UK Biobank, a large-scale, prospective cohort from England, Scotland, and Wales, all-cause mortality increased by 25% for participants with a measured 25-(OH)D concentration of 25 nmol/L compared with 50 nmol/L.

Although this study has a limitation of the lack of information on vitamin D supplementation, a recent Systematic Review and Meta-Analysis of 80 Randomized Clinical Trials found that vitamin D supplementation could reduce CVD events but not CVD or overall mortality. With vitamin D supplementation being recommended for more therapeutic and preventive justification and the high prevalence of vitamin D deficiency in the UAE and the region, its screening and management could be highly recommended to decrease CAD events attributed to it (26, 27). Additionally, debate remains on the best tests for measuring vitamin D and recommended levels of vitamin D intake (28). Optimal levels of circulating 25(OH)D levels vary among countries. Recommended levels based on studies of the musculoskeletal system may or may not be appropriate for other vitamin D-impacted conditions such as immune function, cancer prevention, cardiovascular health, and neurologic function. Only further investigation will settle these issues (28).

Regarding lower GFR levels, there is more consensus on its contribution as a major risk factor for ASCVD (29). Individuals with CKD are more likely to die of a CVD event than to progress to kidney failure. Thomas et al. reported that cardiovascular deaths attributed to reduced GFR outnumbered ESRD deaths throughout the world and that studies are needed to evaluate the benefit of early detection of CKD and treatment to decrease these deaths (30). In fact, GFR is already incorporated in some ASCVD equations, such as the PREVENT equations with glomerular filtration rate as a predictor (31). Matsushita developed “CKD Patche” incorporating eGFR and albuminuria, to enhance the prediction of risk of atherosclerotic CVD (ASCVD) by the Pooled Cohort Equation (PCE) and CVD mortality by Systematic Coronary Risk Evaluation (SCORE). The prediction improved with the CKD Patch for CVD mortality beyond SCORE and ASCVD beyond PCE in validation datasets (32, 33).

An important question raised in this study is the appropriateness of developing one equation for ASCVD instead of two, one for CAD and one for stroke. The definition of ASCVD, which includes CAD and stroke, does suggest that the diseases are related. Analysis of each separately in this study as an outcome, however, yielded different equations. The Framingham study has many equations, with one for hard CAD and one for ASCVD, while the PCE is only for ASCVD events grouped together (8). Further inquiry in this area is vital for more precise patient care.

With the precise causal mechanisms underlying prediction factors remaining unclear, much work remains to be done on interventions that improve health and increase longevity. Access to care was found to be a determinant of longevity and better health outcomes (34). Abu Dhabi has an open-access healthcare system with an average visit to primary care providers of 11 per year, which is very high compared to 3.9 in the US. (35) A further issue is the impact of the Weqaya national screening program on awareness of chronic conditions with the accompanying lifestyle therapeutic interventions directed against diagnosed risk factors, such as lipid-lowering, metformin for prediabetics and new diabetic and weight-lowering medications. This is evident from the results of this study. In Abu Dhabi, there is a 45% lifetime cumulative prevalence of the use of LLM, mainly statins. This is more than reported in the US, for example (17). This study showed not only the high prevalence of LLM use but also their apparent effect on reducing mortality, which added valuable insight to others on cardiovascular risk equations and the influence of therapeutic interventions such as LLMs. A study aiming to determine the effect of statin medication on the predictive capabilities of the PCE in a population-based cohort study showed that including statin prescriptions did not improve risk predictions (36) (37). Nevertheless, there was no study of their influence on overall mortality in relation to the cardiovascular risk assessment. In view of the dynamic nature of such interventions, we suggest developing cardiovascular equations that account for the dynamic nature of risk factors and the effectiveness of such interventions.

Although the COVID-19 pandemic was associated with conditions that likely contributed to the risk associated with increased CAD and stroke mortality, we did not see an increase in mortality in this cohort. In the US, from 2019 to 2020, the increase ranged from 2.3% to 11.9% (1). This was attributed, according to the authors, to complex social determinants involving access to care, poorer medication adherence, and increased barriers to healthy lifestyle behaviors. In Abu Dhabi, there was strong government support to mitigate the pandemic effect in healthcare and other sectors (38).

Worth noting is that some of the risk factors may have been misreported. For example, although smoking was found to be rare among females in this study, underreporting may have occurred as smoking is stigmatized in females in Abu Dhabi. Similarly, race was not included as a studied risk factor in this study. This was not possible as there was no documentation of race. However, this may not be a major limitation, as suggested by Khan et al., as they hypothesized that race factor is affected by social determinants of health and SDOH rather than genetics or ancestry of patients. In calculating the PCE, race was considered nonblack as the vast majority of the UAE origin is not from Africa. Additionally, risk prediction and reduction may be further improved by incorporating a Polygenic Risk Score. There are increasing reports of its role in improving prediction. Marston et al. concluded from a cohort study that the predictive ability of a polygenic CAD was greater in younger individuals and can be used to identify patients with borderline and intermediate clinical risk who should initiate statin therapy (39).

The Kaplan-Meier survival analysis showed a graded risk of mortality based on the developed equation. This can be considered initial validation for another outcome than ASCVD in this population, although more than one-third of deaths are due to CVD in ABU Dhabi, and such a relationship may be expected (2). This performance against the overall mortality rate had a positive linear relation over the follow-up years with the developed hazard ratio. Additionally, such relation was stratified by the impact of therapeutic interventions, the LLMs. Therefore, part of the variation from other international equations could be explained by the high LLM use and not only ethnicity supporting what is currently well noted in the literature, that is, the secular trends in risk factor levels as the decline in lipid levels in the past decade (40).

## Validation

The performance of the ADRS-ASCVD and ADRS-CAD was very good in predicting ASCVD and CAD. In a recent study on the development and validation of the American Heart Association’s PREVENT Equaation, c-statistics in external validation for CVD was 0.794{Khan et al., 2024, #265706}. The change in c-statistic was 0.073 for the ASCVD and 0.10 for CAD nevertheless, both represent good reproducibility. The change in the performance from derivation to validation is common in literature and was attributed to many factors.

A possible factor in this study is the sampling of the cohorts plays an important role, with sampling in the development cohort probably being community-based and more random as subjects were required to enroll in the screening program to get access to healthcare, while in the validation it was community-based as well but completely optional and to increase enrolment intense marketing was done through advertising that comprehensive important investigations are included and that can attract a certain type of population that may be more health-focused or having more symptoms. This is supported by a higher number of events in the validation cohort. As well, the homogeneity of cohorts is difficult to assess and can influence the prediction model performance. This is supported by the fact that the performance of the model was probably not due to lack of fit since the PCE had a similar decrease in the c-statistic of 0.074. Additionally, the difference in the period of follow-up, in the derivation cohort was 9.4 years compared to 6.7 in the validation cohort may have played a role since there is an annual incidence of events, and if included the model performance is expected to be affected. Finally, the change in risk factors is inevitable with the introduction of new medications for hypertension and diabetes, and the use of statins in this cohort at the start of the follow-up was high already, 39.4% compared to the derivation cohort of 14.9%. In a systematic review on the methodological conduct and reporting of studies evaluating the performance of multivariable prediction models (diagnostic and prognostic) there were variations noted between the derivation and validation c-statistics{Collins et al., 2014, #242171}. There was no explanation on the justification for such differences which probably was left for further studies on the difference in areas of studies, study methodology, population, and diseases studied.

The ADRS and the PCE were in agreement on this cohort. This was noticed as well in the development cohort. The CAD formula performed better with 0.7 while the stroke formula performed lowest, 0.7. The smaller events that occurred in the derivation cohort could have affected the accuracy of the formula and more studies focusing on stock can increase the accuracy.

In conclusion, risk assessment is fundamental in the prevention of ASCVD. These results are valuable for Abu Dhabi and the region in risk-stratifying patients for early, effective, individualized primary prevention of CVD. For the Abu Dhabi population, the results of this study will inform patient care and practitioners, as well as future public health measures. External validation is in progress to facilitate its use but this cohort’s unique equation can be informative to others regarding variations that affect its components, proving the dynamic change of risk factors depending on the population, country, and interventions used.

## Declarations

### Ethical approval and consent to participate

The study was approved by the AlAin Human Ethics Committee, approval number 13/58, and Ambulatory Healthcare Services IRB 19-2022. All methods were carried out under relevant guidelines and regulations. The authors confirm that the study was conducted in accordance with the Helsinki Declaration.

### Consent statement in the Ethics approval and consent to participate

Informed consent was waived by the IRBs as the study was designed for retrospective data gathered as part of patient care and anonymized at analysis.

### Authors’ contributions

LBK and NN conceptualized and analyzed data. LBK wrote the manuscript; all other co-authors collected data and reviewed the manuscript. All authors have read and approved the final manuscript.

## Data Availability

Data availability is restricted due to institution policies.

## Acknowledgments

None.

## Consent to publish

Not Applicable.

## Availability of data and materials

Data availability is restricted due to institution policies. Figure 1 Area Under the Curve (AUC), c-statistics, with coordinates of the ROC curve noting the sensitivity and specificity of the equation at different cutoff points. A. The cohort of 8261 CAD-free subjects followed for CAD events over the follow-up period, B. The cohort of stroke-free subjects followed for new stroke development over the follow-up period C. The cohort of 8123 ASCVD-free subjects followed for ASCVD events over the follow-up period. Figure 2 Kaplan-Miere graph of mortality in two strata, on LLM and not on LLM, stratified into ADRS-ASCVD 5 risk levels; <1%, 1-3.99%, 4%-7.49%, 7.5-9.99%, 10-19.9%, and >=20.

For the cohort of 8123 ASCVD-free subjects followed for ASCVD events over the follow-up period. B For the cohort of 8261 CAD-free subjects followed for CAD events over the follow- up period.

