## Appendix 1 for "Cardiovascular disease risk score derivation and validation in Abu Dhabi, United Arab Emirates. Retrospective Cohort Study"

Appendix 1: Follow-up period from screening date to 2023.

|  | Number | % | Cumulative % |
| --- | --- | --- | --- |
| 2011 | 146 | 1.8 | 2.2 |
| 2012 | 195 | 2.4 | 4.5 |
| 2013 | 210 | 2.5 | 7.1 |
| 2014 | 192 | 2.3 | 9.4 |
| 2015 | 171 | 2.1 | 11.5 |
| 2016 | 131 | 1.6 | 13.0 |
| 2017 | 129 | 1.6 | 14.6 |
| 2018 | 150 | 1.8 | 16.4 |
| 2019 | 165 | 2.0 | 18.4 |
| 2020 | 254 | 3.1 | 21.5 |
| 2021 | 655 | 7.9 | 29.4 |
| 2022 | 1861 | 22.5 | 52.0 |
| 2023 | 3968 | 48.0 | 100.0 |
| Missing | 229 | 100 |  |
| Total | 8456 |  |  |
