## Appendix 2 for "Cardiovascular disease risk score derivation and validation in Abu Dhabi, United Arab Emirates. Retrospective Cohort Study"

ASCVD

COMPUTE ASCVDFormula=((VITD_VALUE- 35.53) * (- 0.007))+ ((deviationGFR- 0) * (-0.029))+ ((SEX-

0.51) * 1.516) + ((CurrentSmoker- 0.09) * 0.611) + ((Age - 38.6) *0.204 ) + ((BPtreated - 0.13)

*0.305 ) + ((Chole_HDL_ratio- 4.10) * 0.068) + ((DMDx - 0.21) * 0.332) + ((deviationGFRSEX -

(-1.28)) * 0.031) + ((mean_BP - 89.32) * 0.012) + ((AGE_SQR- 1749.50) * -0.001).

EXECUTE.

COMPUTE ASCVD_SUMRF_=EXP(ASCVDFormula).

EXECUTE.

COMPUTE ASCVD_SUMRF_risk=1 - (0.994 ** ASCVD_SUMRF).

EXECUTE.

COMPUTE ADRS_ASCVD_SUMRF_risk=ASCVD_SUMRF_risk * 100.

EXECUTE.

CAD

COMPUTE SUMRF=((VITD_VALUE - 35.53) * -0.007) + ((deviationGFR- 0) * -0.039) + ((SEX- 0.51) * 2.136) + ((CurrentSmoker- 0.09) * 0.55) + ((Age - 38.6)

*0.067 ) + ((Hypertension - 0.18) *0.327 ) + ((chole_HDL_ratio - 4.1) * 0.067) + ((DMDx - 0.21) * 1.543) + ((deviationGFRSEX - (-1.28)) * 0.027) +

((DMSEX - 0.12) * -0.669) + ((DMdevGFR- (-2.72)) * 0.019) + ((BP1 - 0.46) * -1.118) + ((BP2 - 0.15) * -1.148) + ((BP3- 0.25) * -1.378) + ((BP4 - 0.14) * -1.093) + ((BP5 -

0) * -0.698).

EXECUTE.

COMPUTE XPSUMRF=EXP(SUMRF).

EXECUTE.

COMPUTE RISK_XPSUMRF=1 - (0.994 ** XPSUMRF).

EXECUTE.

COMPUTE PRCENT_RISK_XPSUMRF=RISK_XPSUMRF * 100.

EXECUTE.

Stroke

COMPUTE SUMRFMM_STRK=( CurrentSmokm * 1.053)+(SBP_mean * 0.024)+ (Agemean * 0.084)+(HTNSEX_mean * -0.646) + (AGESEX_mean * 0.011).

EXECUTE.

COMPUTE sumrfFSTRK=( CurrentSmoker * 1.053)+(SBP * 0.024)+ (Age * 0.084)+(HTNSEX * -0.646) + (AGESEX * 0.011).

EXECUTE.

COMPUTE sumrfFSTRK=( CurrentSmoker * 1.053)+(SBP * 0.024)+ (Age * 0.084)+ (AGESEX * 0.011).

EXECUTE.

COMPUTE SUMRFF_SUMMM_STRK=sumrfFSTRK - 6.37.

EXECUTE.

COMPUTE XP_SUMRFF_SUMMMSTRK= EXP (SUMRFF_SUMMM_STRK).

EXECUTE.

COMPUTE PROP_RISKSTROKE=1 - (0.998 ** XP_SUMRFF_SUMMMSTRK).

EXECUTE.

COMPUTE STROKE_RISK_PERCENT=PROP_RISKSTROKE * 100.

EXECUTE.
