## Appendix 3 for "Cardiovascular disease risk score derivation and validation in Abu Dhabi, United Arab Emirates. Retrospective Cohort Study"

Appendix 3 The stratification by risk categories based on the ADRS formula distributed by the occurrence of an ASCVD event during the follow-up period. In the derivation cohort in A, B, and C, and the validation cohort in D.

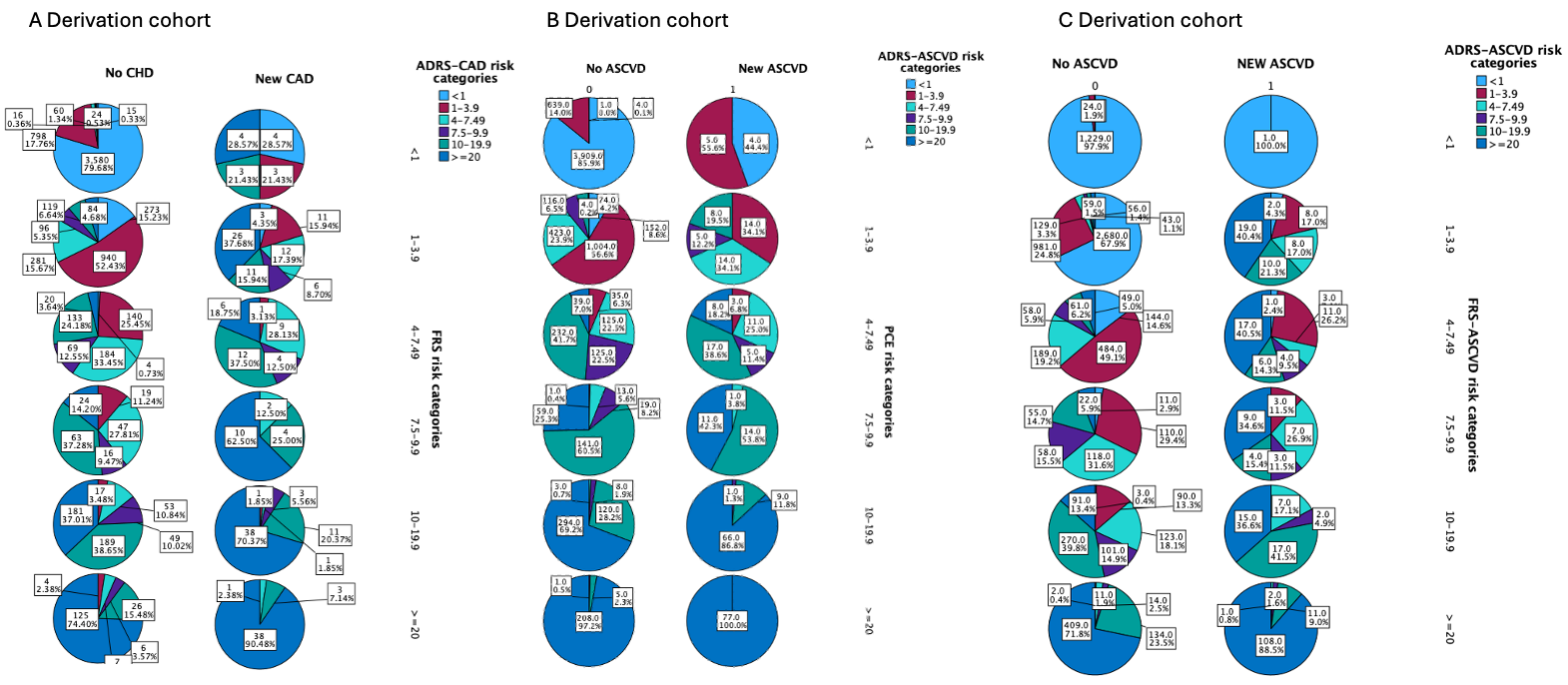

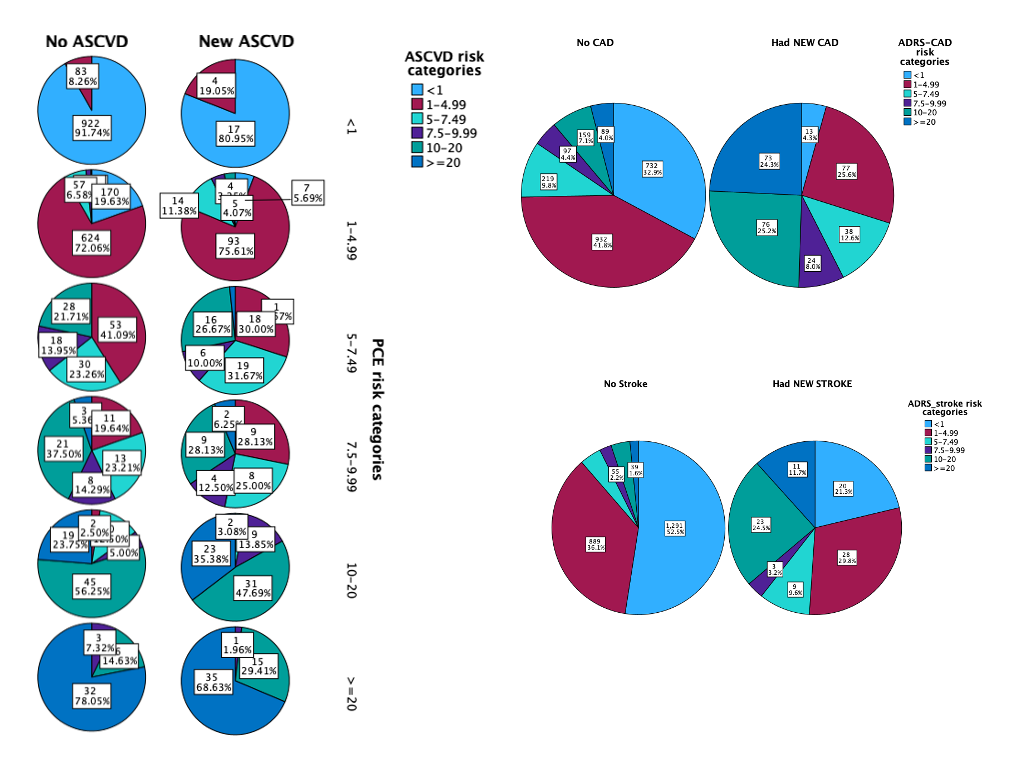

D Validation cohort
